# Effects of primary-care led self-management and low carbohydrate diet on glycaemic control in individuals with type 2 diabetes mellitus: study protocol of a cluster randomised controlled trial

**DOI:** 10.1101/2024.11.23.24317824

**Authors:** Joshua Chadwick, P Ganeshkumar, Jeyashree Kathiresan, Hemant Deepak Shewade, K Madhanraj, S Devika, S Lokesh

**Author notes:** **Corresponding author** Dr Joshua Chadwick, Scientist-B (Medical), ICMR-National Institute of Epidemiology, R-127, TNHB, Ayapakkam, Chennai, Tamil Nadu, India 600077, Ph: Off+91 44 26136208.

## Abstract

**Introduction:** Type 2 diabetes mellitus (T2D) poses a significant global health burden, contributing to mortality and morbidity primarily through its complications such as renal failure, cardiovascular events, and others. Dietary patterns, particularly low carbohydrate diet and diabetes self-management education (DSME) have shown promise in improving glycaemic control and other metabolic parameters. However, evidence on the safety and long-term efficacy of such diets including health education, especially in populations like India with a high carbohydrate dietary tradition, is limited. This paper outlines a randomised controlled trial to determine the effectiveness of adopting a low carbohydrate diet in addition to standard care for T2D patients compared to those following standard care alone.

**Methods and analysis:** This is a cluster randomized controlled trial that adheres to the Recommendations for Interventional Trials as outlined by the Standard Protocol Items. The study will be conducted in 16 urban primary health centres (UPHCs). UPHCs randomized to either standard care or low carbohydrate diet intervention. Outcome: generating evidence on improving HbA1c levels without increasing anti-hyperglycaemic medication. This will be achieved through the use of CGM technology and personalized nutrition counselling, which will focus on dietary carbohydrate restriction for patients with T2DM.

**Discussion:** Results of this study will be published in peer-reviewed journals. The trial will provide insights into the efficacy and feasibility of implementing low carbohydrate diets and DSME in T2D management within public health settings. This protocol is registered on Clinical Trials Registry-India. The registration number for this trial is CTRI/2024/02/062202.

## Introduction

With an estimated 1.5 million deaths primarily due to diabetes in 2019, diabetes was the ninth most leading cause of mortality.[1] Compared to high-income countries, prevalence has been increasing more rapidly in low- and middle-income nations.[2] In India, the prevalence of diabetes was 7.3%.[3] The World Health Organization (WHO) has identified unhealthy diets and a lack of exercise as risk factors for non-communicable diseases.[4]

Systematic reviews and meta-analyses of randomised control trials (RCTs) suggests low carbohydrate diets in T2D may enhance glycaemic control, triglyceride levels, and HDL cholesterol profiles.[5–8] In 2020, American Diabetes Association (ADA) standards of care for Type 2 Diabetes (T2D) report stated: ‘for individuals with type 2 diabetes not meeting glycaemic targets or for whom reducing glucose-lowering drugs is a priority, reducing overall carbohydrate intake with a low-carbohydrate or a very-low-carbohydrate eating pattern is a viable option’.[9]

Indian diet has a tradition of relying primarily on carbohydrates, making it increasingly challenging to control diabetes.[10] Indians typically consume around 65% of calories though carbohydrates and are heavily dependent on their staple grains’ consumption.[11] However, little is known about the most practical ways to put recommendations on lower carbohydrate diets into practice in public primary care settings in India.

### Objectives

The primary objective of the study is to evaluate the effectiveness of intervention model for low carbohydrate dietary adherence and diabetes self-management in individuals with type 2 diabetes mellitus, aiming to enhance the capability for self-management and adherence to a low carbohydrate diet, especially the value of glycated haemoglobin (HbA1c), within the routine primary care setting where individuals with T2D is manged. The secondary objectives are: a) to evaluate the cardio-renal risk variables such as body weight, lipid profile, serum creatinine, blood pressure (BP), homeostatic model assessment for insulin resistance (HOMA IR), effect on diabetes medication prescription and change in the monthly cost of antihyperglycemic agents and any adverse events, b) To describe the meal-related glycaemic fluctuations based on real-time continuous glucose monitoring tool, c) to determine the health-related quality of life (HRQoL) using the global health visual analogue scale (EQ-VHA) and the EuroQol five-dimensional questionnaire (EQ-5D), and d) to evaluate the enablers and barriers of adapting self-management practices and low carbohydrate diet from the perspectives of individual and service provider.

## Methods

### Study design

This trial is conducted in accordance with the Standard Protocol Items: Recommendations for Interventional Trials (SPIRIT) as a cluster randomized controlled trial. Recruitment of patients for this yet-to-begin study is scheduled to commence in November 2024. The study will use a cluster-randomised controlled design with a nested mixed-methods process evaluation, with urban primary centres (UPHC) as the unit of randomisation. Randomisation at the UPHC level will prevent contamination between groups and allow consistent advice from project assistant, who will have a background in nursing or life sciences, who will be offering the self-management and dietary advice.

The Indian Council of Medical Research (ICMR)-National Institute of Epidemiology, Chennai, will centrally and independently of the study team conduct the zone randomization and the selection of the UPHC (Figure 1). The protocol was reviewed and approved by the Scientific Advisory Committee/ Institutional Human Ethics Committee of the research institution (NIE/IHEC/202302-03) and also approval was obtained from the corporation health officer, Greater Chennai Corporation (P.H.D.No.C6/06658/2023). The study has been registered in the Indian Clinical Trial Registry (CTRI/2024/02/062202).

**Figure 1.**
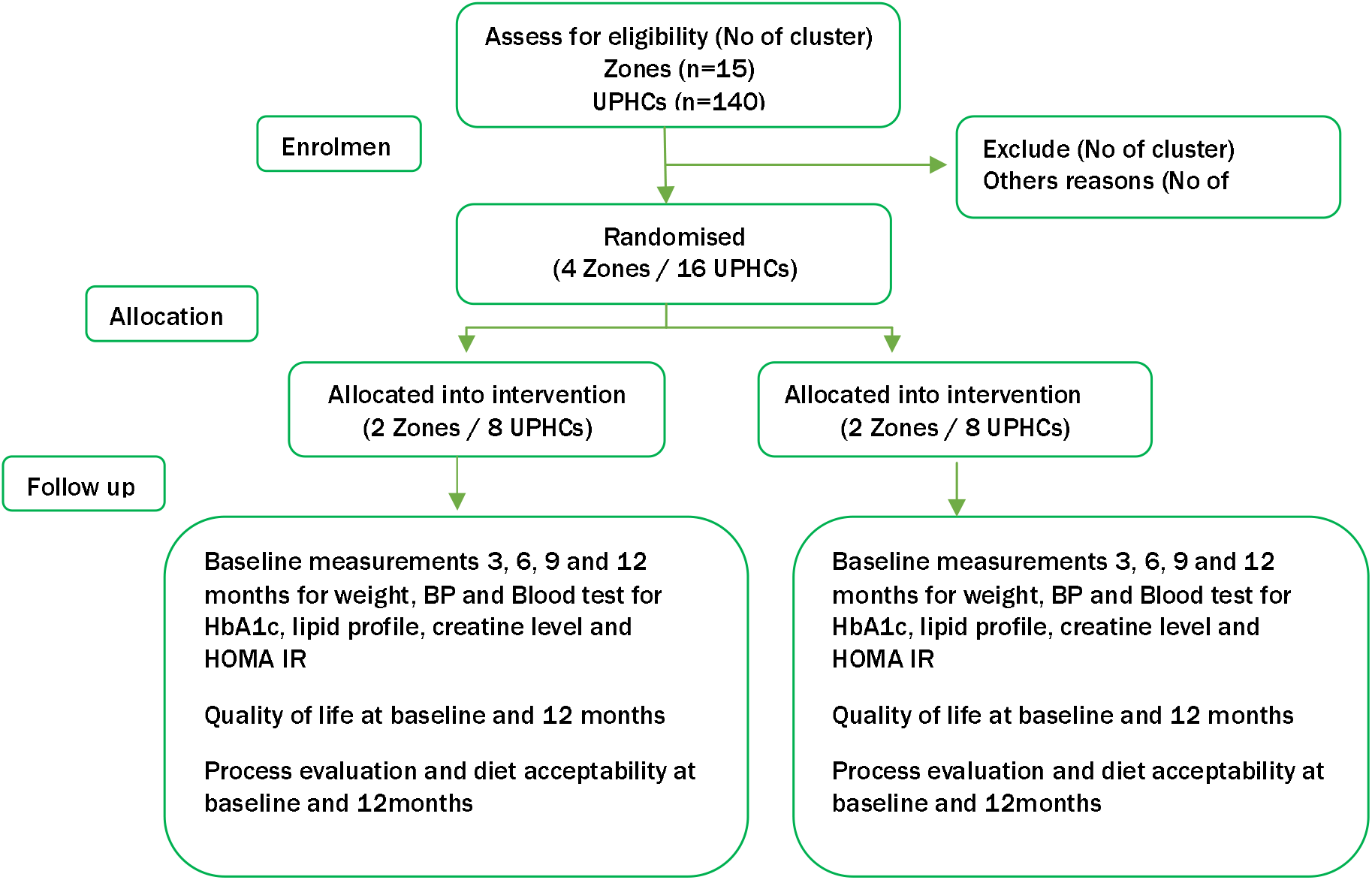
Depicting the CONSORT flow chart of the trial.

### Study setting

The study will be conducted at UPHCs of Greater Chennai City Corporation, South India.

### Targeted population

Individuals with type 2 diabetes who is seeking health care in UPHCs in Greater Chennai city corporation.

### Inclusion and exclusion criteria

Individuals over the age of 30 years who have type 2 diabetes with or without comorbid conditions, HbA1c level that is greater than 7.5% during the screening test will be included in this study and those who having terminal illness, mental illness, learning difficulties, diagnosed eating disorder or purging, pregnant/ considering pregnancy in the next 12 months and current use of weight-loss drugs or use of oral glucocorticoids will be excluded.

### Recruitment

A total of 320 participants (160 each arm) will be recruited from the selected UPHCs (figure 1). Participants who meet the criteria will be included. Reasons for exclusions will be recorded. Informed consent will be secured from all participants, for 18 months on the trial protocol and indefinite long-term data collection from their medical records and through population health registry.

Individual participants will be randomly assigned in a 1:1 ratio to either the intervention or control group. We will ensure that all participants have an equal chance of being allocated to either group to minimize selection bias. To limit attrition and analytical bias, we will use intention-to-treat (ITT) analysis.

### Blinding

It will not be possible to blind the investigators or participants to allocation. Those responsible for analysis will be blinded to allocation.

### Intervention

Participants in the intervention arm (Zone/UPHCs) of the study will be provided with advice on adopting a low carbohydrate diet (<130 g of carbohydrates/day) and health education for diabetes self-management, in addition to receiving standard medical care. During their regular physician visits, participants will receive dietary advice for a period of 10-20 minutes. They will be recommended to follow a low carbohydrate diet and health education, and a one-on-one visit will be scheduled every 3, 6, 9, and 12 months to monitor their progress. Monthly telephonic follow-ups will also be conducted to track their dietary habits. For the first eight weeks, patients will be monitored closely for any signs of hypoglycaemic symptoms, and their blood glucose levels will be checked as required. The control arm will be advised to follow a standard diet, along with the standard diabetes treatment. They will have 10-20-minute ‘one-on-one’ visits for advice on following a standard diet at baseline, 3, 6, 9, and 12 months.

During the initial two weeks, a total of 30 individuals (15 in intervention and control arm) with T2DM will have the opportunity to wear a CGM and keep a log of their diet, hypoglycaemic symptoms, and blood glucose level, which will be checked whenever required. Strategies to improve adherence will be implemented at every visit, including counselling on the importance of adhering to the prescribed diet and medication, as well as guidance on what to do in the event of hyperglycemic episodes.

Patients will also be instructed on the purpose and use of the study diet. Challenges encountered during the study will be discussed at each visit, and patients will be informed about contacting the study centre or study physician for any difficulties in following the diet or experiencing symptoms of hyperglycaemia. Patients who experience drug intolerance, diet intolerance, or poor compliance will be documented and included in the intention-to-treat analysis.

The Diabetes Plate Method handouts will be used to instruct participants on how to divide the plate. Infographic of balance between sugar intake and output also will be given to study participants (Figure 2). The lower carbohydrate diet sheet will be given which provides a sample diet chart and information on low GI food sources (annex 1).

Information on the glycaemic load derived from the glycaemic Index (GI) will be tailored to meet the sociocultural dietary preferences of each participant. This approach aims to encourage a reduction in the consumption of foods high in sugar and starch, such as rice-based dishes like dosa or idly, rice, or bread, and to suggest alternatives including green leafy vegetables, legumes, millets, full-fat dairy products, eggs, meat, fish, guava, berries, and nuts. By doing so, it will equip participants with the knowledge to make healthier food choices by comparing the glycaemic load of different meal options to the glycaemic impact of a standard amount (4 grams) of table sugar per teaspoon.[13]

### Sample Size

Evidence from the literature showed that changes in the HbA1c level for three months is 0.6 in the Low-Carbohydrate Diet group and 0.3 in the standard diet with the standard deviation of 1.2 and 0.7, respectively.[14] Therefore, we calculated 85 patients for each group, with α = 0.05 and power = 0.80. In view of the sample loss of 20% and design effect of 1.5, the number for each group is 153 rounded to 160. Assuming 20 subjects to be enrolled in each UPHC we need 8 UPHC from 2 Zones for each group.

### Data collection, management, and analysis

#### Baseline assessments and subsequent visits

Baseline measurements of weight and BP along with blood tests such as HbA1c, lipid profiles, creatinine level and HOMA IR. Follow-up will be for a year at 3, 6, 9 and 12 months. During the follow-up visits, variables such as body weight, HBA1c levels, BP, lipid profiles and creatinine will be measured. Change in the number of antihyperglycemic medications will be determined by subtracting the total number of antihyperglycemic medications at baseline from the total cost of antihyperglycemic medications during follow-ups.

#### Diet acceptability, process evaluation and quality of Life

Semi-structured questionnaires will be used to gather information from study participants and their family members through face-to-face interviews at the UPHC or telephone interviews. The questionnaires will focus on their experiences, challenges, facilitators, and strategies for adapting to a low-carbohydrate diet till data saturation is achieved.[15] The intervention group participants will be interviewed three times: once after the use of CGM (continuous glucose monitoring), at months 1 and 12. The control group participants will be interviewed twice, at months 1 and 12. The interviews will be conducted by team members who are trained in semi-structured interviewing techniques, and they will receive regular feedback from a qualitative methodologist. The implementation of the intervention and the ease of engagement will be assessed by a purposive convenience sample of 10 health care professionals (doctors, nurses, and dietitians) who will be providing the intervention, will be interviewed. Quality of Life outcome (EQ-5D-3 L) will be measured at baseline, 6 and 12 months for all participants.

#### Statistical methods

Descriptive analysis will be performed for all the quantitative variables and the results will be summarised as proportions and mean ± SD. The primary outcomes, which include changes in HbA1c levels, blood pressure, lipid profiles, creatinine, and body weight between groups, will be analysed using a paired t-test. If the participants withdraw from the intervention will be included in ITT analysis. Additionally, a thematic analysis will be carried out on the qualitative data.

#### Monitoring

The trail will be supervised by data safety and monitoring board (DSMB). The board composed of 5 members independent of the Investigators, having expertise in diabetes, low carbohydrate diet, nutrition and ethics. DSMB will review the study at predetermined intervals and issue recommendations concerning ongoing study in order to ensure that risks are minimized and benefits are maximised for patients and also the Investigator will ensure that this study is conducted in accordance with Good Clinical Practice.

### Data management

The ODK app will be used for quantitative data collection, which will be transferred to the ICMR-NIE server. Qualitative data will be collected through an audio recorder after obtaining consent.

### Ethics and dissemination

The Investigator will ensure that this study is conducted in accordance with the principles of the Declaration of Helsinki. The protocol, informed consent form, participant information sheet, any participant facing materials and infographic material has been submitted to Institutional Human Ethics Committee (IHEC) and/or regulatory authorities and have sought the written approval (NIE/IHEC/202302-03).

### Consent

A written informed consent shall be obtained from the participant. In the event that the participant or their relative lacks formal education, a thumbprint shall be taken. The Participant Information Sheet (PIS) has been made available in English and Tamil shall provide comprehensive information regarding the study’s nature, benefits, implications, limitations, side effects, and risks. The participant shall be free to withdraw their consent or discontinue participation at any time, without any prejudice to future care, and without any obligation to provide a reason.

### Confidentiality

The study team will ensure that participants’ anonymity will be maintained and their records will be kept confidential. All documents related to the study will be securely stored and accessed only by authorized personnel and the study staff. Patient names and any identifying information will not be entered into any digital file or document, including the eCRF.

### Dissemination policy

Findings will be shared with competent authorities such as the Greater Chennai City Corporation and their UPHCs, and will be published in high-impact journals, in accordance with CONSORT guidelines.

## Discussion

The management of T2D continues to be a significant global health challenge, particularly in low- and middle-income countries like India. Despite advancements in therapeutic options, glycaemic control in patients remains suboptimal, often due to various socio-cultural, economic, and dietary factors. This cluster-randomised controlled trial aims to address this challenge by examining the effects of a low-carbohydrate diet and diabetes self-management education (DSME) on glycaemic control in individuals with T2D within the primary healthcare setting of Chennai, India. This study seeks to bridge this gap by providing culturally tailored dietary advice tha[16] aligns with the carbohydrate consumption patterns of the Indian population.[16]

An important strength of this study is the use of real-time continuous glucose monitoring (CGM) to capture meal-related glycaemic fluctuations, which will provide objective data on the effectiveness of dietary interventions in controlling postprandial hyperglycaemia. Furthermore, the study design includes a robust process evaluation component, which will assess both the fidelity and feasibility of the intervention. This approach ensures that the findings will not only focus on clinical efficacy but also on the practicalities of delivering the intervention in routine care.

One potential limitation of trial we foresee is attrition, which we aim to minimize through regular follow-up, telephone consultations, and an ITT analysis.

If successful, the findings from this trial will have significant implications for diabetes management, particularly in low-resource settings. The study will offer evidence on how dietary interventions, combined with DSME, can be integrated into primary healthcare systems in India, providing a low-cost, scalable approach to improving glycaemic control. Moreover, the outcomes of this study will be crucial in informing future guidelines and policies on dietary interventions for T2D management, particularly in populations with traditionally carbohydrate-rich diets.[17,18]

In conclusion, this trial represents an important step toward optimizing diabetes care through dietary modifications and self-management education. The results of this study will provide valuable insights into the feasibility, safety, and effectiveness of low-carbohydrate diets in controlling T2D, contributing to the growing body of evidence that supports individualized, culturally appropriate dietary interventions in the management of chronic diseases.

## Data Availability

All data produced in the present study are available upon reasonable request to the authors

## Authors Contributions

This is a collaborative research, each of the listed people provided important intellectual contribution to study design. JC drafted the protocol. JC, SL, PG, JK, HDS and MK drafted the final manuscript, while JC revised the final protocol. All authors contributed to data analysis, drafting and revising the article, and have read and agreed to the published version of the manuscript.

## Patient and public involvement

Patients and/or the public were not involved in the design, or conduct, or reporting, or dissemination plans of this research.

---

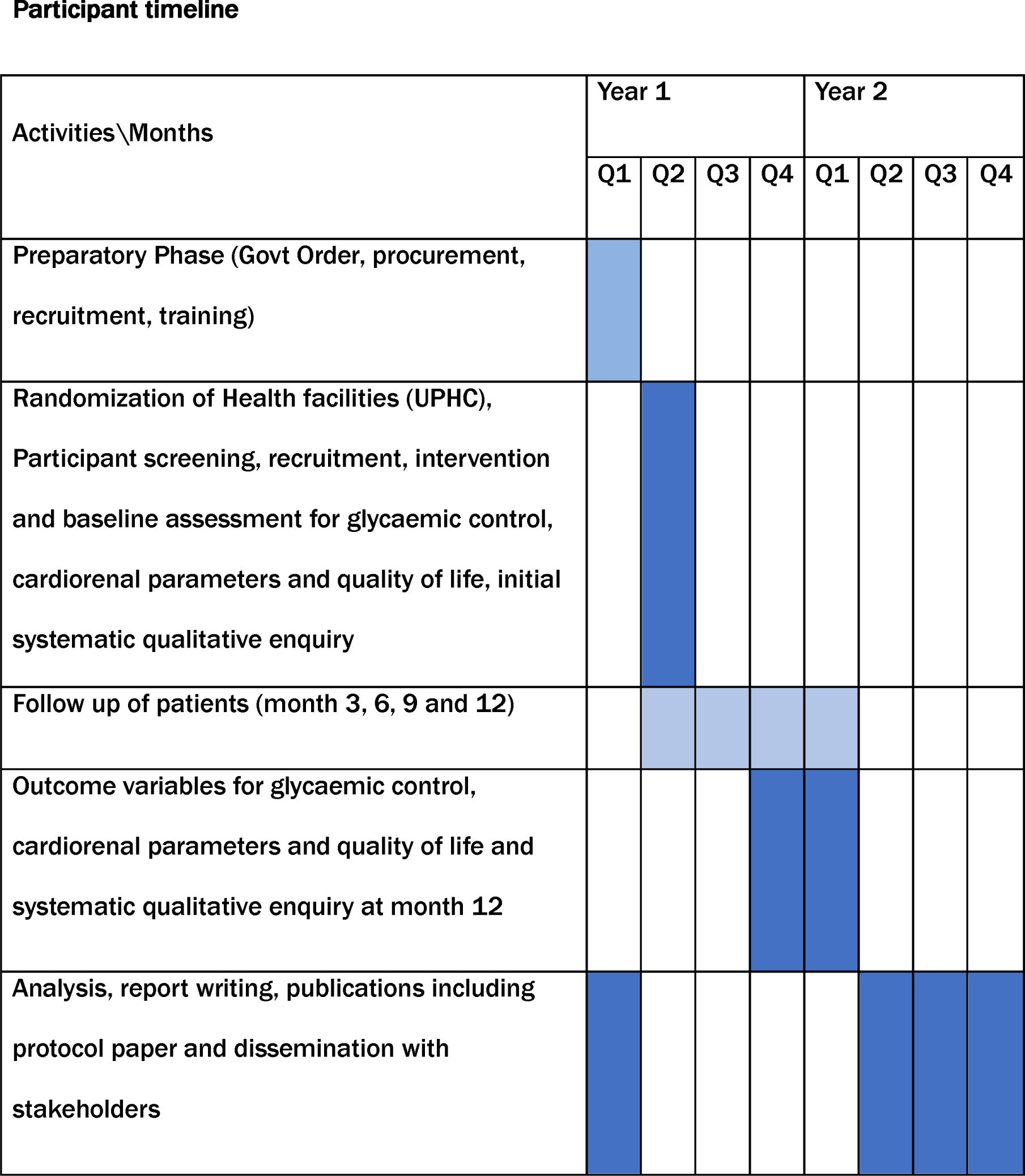

